# Psychological factors contribute more to chronic low back pain than spine pathology: An LLM-based analysis of radiology reports

**DOI:** 10.64898/2026.09.16.26363055

**Authors:** Mattia Perrone, Hiroyuki Tachi, Hanadi A. Albadi, D’Mar M. Moore, Ashrith V. Alavilli, Nathan J. Lee, Tricia J. Johnson, John T. Martin

## Abstract

**Background:** Chronic low back pain (cLBP) is the leading global cause of disability, yet anatomical, psychological, and socioeconomic determinants are typically studied in isolation, with limited large-scale integration of imaging and biopsychosocial data. We linked imaging findings with demographic, anthropometric, and biopsychosocial characteristics to determine their relative contributions to cLBP.

**Methods:** We retrospectively analyzed 1,480 Chicago-based patients with lumbar spine MRI between 3/2014 and 3/2024. cLBP was as two or more LBP diagnoses recorded six months apart. Spinal pathologies (stenosis, disc disease, facet arthropathy and spondylolisthesis) were extracted from radiology reports using a large language model (GPT-4) and benchmarked against a regex (rule-based text search) pipeline. Multivariable logistic regression evaluated associations between cLBP diagnosis and demographic, anthropometric, socioeconomic, psychological, and anatomical determinants. A parallel linear regression modeled overall anatomical pathology burden.

**Results:** The LLM pipeline achieved an F1-score of 0.98 versus 0.92 for regex. cLBP diagnosis was most strongly associated with depression (OR=2.69, p<0.001) and anxiety (OR=2.31, p<0.001), with smaller contributions from spinal stenosis (OR=1.07, p=0.006) and spondylolisthesis (OR=1.15, p=0.039). Leave-one-domain-out analysis showed psychological factors were most impactful (ΔAIC=120.1), followed by anatomical (ΔAIC=37.0), demographic/anthropometric (ΔAIC=21.8), and socioeconomic (ΔAIC=3.0). Overall anatomical burden was associated with age, male sex, higher BMI, and Non-Hispanic White race, with no associations with socioeconomic or psychological variables.

**Conclusions:** Factors associated with diagnosed cLBP and those associated with structural spinal pathology are largely dissociated. Psychological comorbidities show the strongest associations with cLBP, while anatomical burden is mostly driven by demographics, reinforcing the psychosocial nature of cLBP.

## Introduction

Low back pain (LBP) is the world’s leading cause of disability (1). It is a driver of extreme healthcare utilization, decreased productivity, reduced quality of life and a critical public health burden (2). While most LBP will spontaneously resolve, 10% of individuals develop chronic LBP (cLBP), which no clear clinical treatments can improve (3) (4). The causes responsible for the onset of cLBP are complex and influenced by many factors (5), and despite significant public investment, research effort, and clinical effort, there has been minimal improvement in patient outcomes. cLBP is a heterogeneous disease with underlying socioeconomic, psychological and anatomical drivers that are typically investigated independently of each other. One bottleneck is the inability to comprehensively analyze spine disease at scale to determine psychological and social correlates due to the costly and time-consuming process of acquiring and analyzing spine images. A clearer understanding of how these determinants interact is crucial to improve clinical assessment and guide targeted interventions (6).

At a population level, integrating imaging findings into a biopsychosocial framework requires a scalable approach that can extract meaningful imaging information from routine care. Radiology reports are a widely available description of patient-level spinal pathology, reflecting how imaging findings are interpreted in clinical practice (7) (8). Prior studies have demonstrated the feasibility of extracting pathologies from free-text radiology reports using rule-based or machine learning-based natural language processing approaches, with machine learning methods often providing improved performance and scalability (9) (10) (11). However, the relationship between imaging findings and LBP remains unclear and controversial. Reported associations between LBP and common degenerative findings, including disc pathologies, modic changes, spinal stenosis, and spondylolisthesis, have been inconsistent across studies, with some reporting significant associations (12) (13) (14) (15) and others weak or absent ones (5) (16) (17) (18) (19) (20). Additionally, existing work has not integrated image-based metrics of spinal pathology with patient demographic and anthropometric characteristics, socioeconomic and psychological determinants of LBP in a unified framework. While imaging remains essential for characterizing spinal pathology, LBP is multifactorial and interactions between social context, mental health, and structural pathology may influence symptom presence beyond the contribution of any single domain (5).

Previous work reinforces the biopsychosocial model of LBP and highlights the importance of a broad set of covariates in a comprehensive analysis of LBP. Demographic characteristics such as age, sex, ethnicity and anthropometric factors like body mass index (BMI) are typically incorporated as baseline factors in epidemiological studies related to LBP (21), (22), (23) (24) (25) (26) (27) (28). Socioeconomic factors such as neighborhood income and educational attainment are commonly analyzed as determinants of LBP, in part because they can be derived from patient address and publicly available data (29) (30). Community-level socioeconomic metrics similarly drive many chronic diseases, where higher levels of segregation and socioeconomic inequality may amplify the association between socioeconomic factors and LBP (31) (32) (33). Psychological factors, particularly depression and anxiety, predicts both the onset and persistence of LBP (34) (35) (36), with a stronger influence on pain severity than imaging findings of degenerative spine disease (37) (39) (40), perhaps due to a bidirectional relationship between psychological factors and LBP (41) (42).

Integrating these characteristics into a unified framework requires a large dataset where each segment of a heterogeneous population is well-represented. Chicago, IL, USA is among the most racially and socioeconomically segregated cities in the United States with many underlying health disparities. Increased rates of chronic disease contribute to a life expectancy gap of up to 20 years in under-resourced Chicago communities (43). Thus, Chicago is an ideal setting to evaluate chronic spine diseases in the context of psychosocial drivers of health. The objective of this study was to quantify the relative contributions of patient demographic and anthropometric characteristics and socioeconomic, psychological and anatomical determinants to the presence of LBP in a unified framework. To do so, we extracted patient data from Rush University System for Health (RUSH), a tertiary and quaternary academic health system serving the metropolitan area of Chicago. To enable the inclusion of anatomical determinants, spinal pathologies were mined from free-text lumbar spine radiology reports using natural language processing, comparing the performance of pattern-based text matching with that of a large language model. To contextualize the role of anatomical findings within the broader biopsychosocial framework, we additionally evaluated the associations between spinal pathologies and patient characteristics, socioeconomic and psychological factors.

## 2. Methods

### 2.1 Study design and population

This study was designed as a retrospective observational analysis using electronic health record (EHR) data from Rush University System for Health (RUSH), a tertiary and quaternary academic health system that includes three hospitals (RUSH includes Rush University Medical Center, Rush Copley Medical Center and Rush Oak Park Hospital), serving the metropolitan area of Chicago, IL, USA. Patients living in Chicago, confirmed by residential ZIP code, that underwent lumbar spine magnetic resonance imaging (MRI) between March 2014 and March 2024 were identified by querying records using the corresponding CPT code (72148, 72158) **(Figure 1)**. The study protocol was reviewed and approved by the Rush University Medical Center Institutional Review Board. An initial cohort of 4,671 patients with lumbar spine MRI examinations was identified. The cohort was further limited to patients with at least one RUSH primary care visit within the two years prior to the index MRI. To further isolate non-specific chronic LBP, we excluded those with active neurological conditions at the time of MRI using ICD-10 codes for radiculopathy (M54.16, M54.17), sciatica (M54.30, M54.31, M54.32), neurogenic claudication (M48.06, M48.07) and polyneuropathy (G62.9). To enable the assessment of LBP chronicity, patients were restricted to those that had at least two visits more than six months apart with an MRI between the two visits. The final cohort consisted of 1,480 patients.

**Figure 1:**
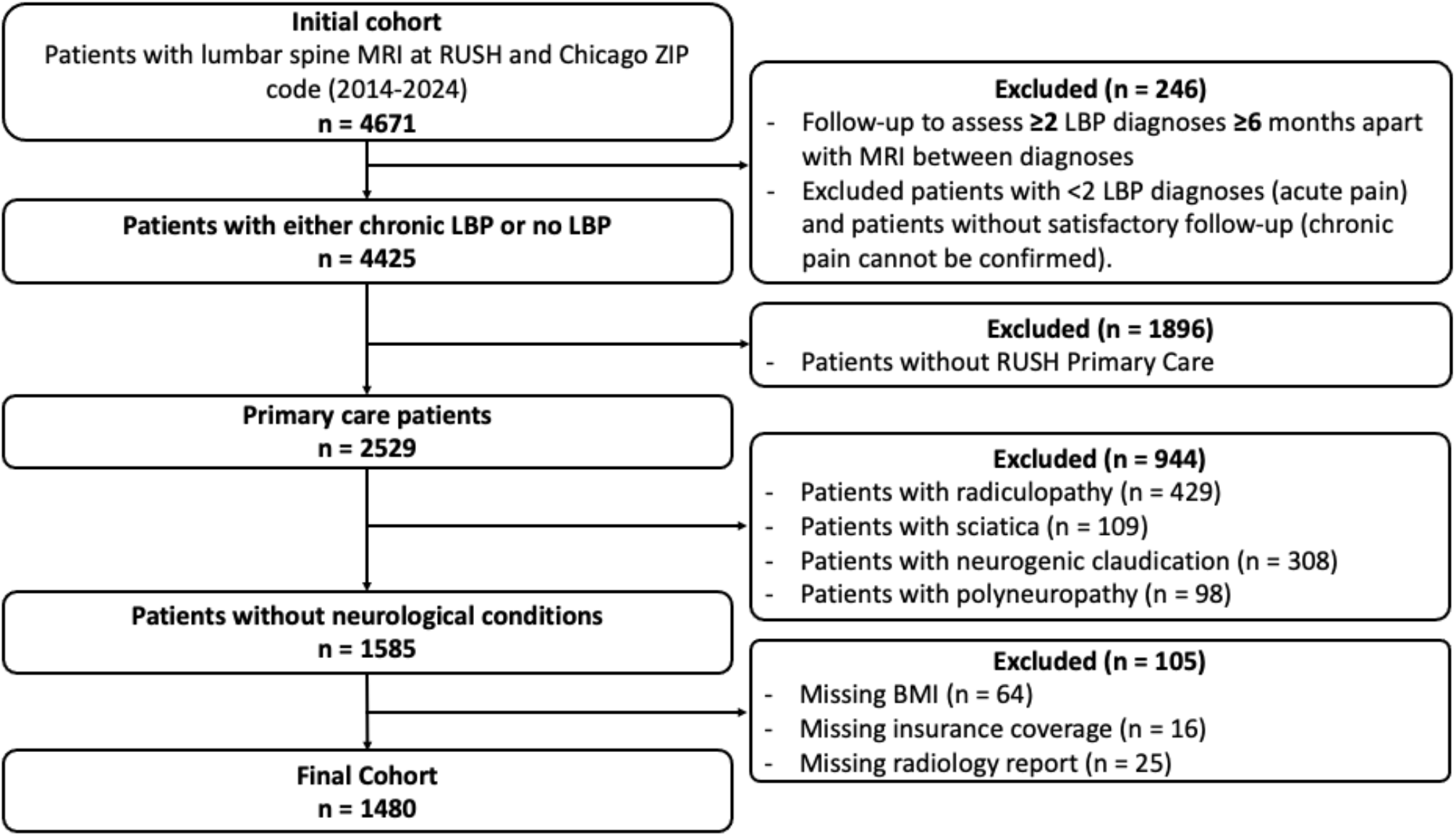
Flowchart showing the application of inclusion and exclusion criteria used to derive the final cohort of patients undergoing lumbar spine MRI.

### 2.2 Outcome

Chronic low back pain (cLBP) was defined using ICD-10 diagnosis codes (M54.40, M54.41, M54.42, M54.49, M54.50) and modeled as a binary outcome. Patients were classified as having cLBP if two or more LBP diagnoses were recorded at least six months apart with an MRI between the two visits. To account for the multifactorial nature of LBP, determinants from four domains were examined: demographic and anthropometric, socioeconomic, psychological, and anatomical.

### 2.3 Determinants

#### 2.3.1 Demographics and anthropometrics

Demographic determinants included age, sex, body mass index (BMI), and race/ethnicity. Age and BMI were modeled as continuous variables, while sex and race/ethnicity were treated as categorical. Race and ethnicity were combined into a single mutually exclusive variable with four categories reflecting Chicago demographics: non-Hispanic Black, non-Hispanic White, Hispanic and other racial groups. Patients with missing BMI were excluded from the analysis (n=64).

#### 2.3.2 Socioeconomic factors

Health insurance at the time of the index MRI visit was used to assess individual socioeconomic status. Health insurance was obtained from structured electronic health record fields and treated as a categorical variable, categorized as private or public. Patients without insurance coverage were excluded (n=16). Community-level socioeconomic factors included census tract-level measures of income and education. Median household income and college graduation rate were derived at the census tract level by linking patient residential addresses to publicly available census data (44) and were modeled as continuous variables.

#### 2.3.3 Psychological factors

Psychological determinants included diagnoses of depression related conditions (ICD-10 code: F32.x, F33.x) and anxiety related conditions (ICD-10 code: F41.x and F43.x), and they were modelled as binary variables. Diagnoses occurred within +/- 15 days of the lumbar spine MRI regardless of encounter type (45).

#### 2.3.4 Anatomical factors

Anatomical information was derived from lumbar spine MRI radiology reports using a commercially available large language model (LLM; GPT-4, OpenAI) accessed via an application programming interface (API). The LLM was used to generate structured representations of radiology findings by spinal level, accounting for variability in terminology and negation. This extraction pipeline was guided by a prompt that instructed the LLM to identify only positive spinal pathologies described in radiology reports, excluding negated or absent findings. The prompt constrained outputs to a predefined set of pathology labels, mapped synonyms to standardized categories and required structured output specifying pathology type, severity (e.g. mild, moderate, severe) when available, and spinal level. Prior to processing, all reports were deidentified to remove protected health information. Patients without radiology reports were excluded (n=25). We benchmarked the performance of the LLM against a natural language processing (NLP) approach based on a rule-based text search using regular expressions (regex). Regex rules were designed to identify predefined keywords corresponding to specific spinal pathologies using pattern-matching techniques. This approach accounted for lexical variations (e.g. disc bulge, disc bulging, bulged disc) to improve pathology detection. Extraction performance was evaluated on a manually annotated set of 100 radiology reports, with LLM-based and regex-based approaches compared directly against manual review

Spinal pathologies extracted from radiology reports included four anatomical factors: disc related pathologies (disc bulging and herniation, narrowing, desiccation, degeneration), spinal stenosis (central canal, foraminal, lateral recess), facet arthropathy and listhesis (anterolisthesis and retrolisthesis). When severity descriptors were reported (e.g. mild, moderate, severe), these were recorded and incorporated into a composite anatomical severity score reflecting both the number of affected levels and reported severity, with higher weights assigned to more severe findings and to pathologies involving multiple levels. Severity grades were assigned at each lumbar vertebral level (L1-L2 through L5-S1) using a 3 point scale: 0 (absent), 1 (mild-moderate), 2 (severe). Mild and moderate categories were collapsed to minimize subjective interpretation across radiologists’ reporting styles. When severity descriptors were not reported, the mild-moderate category was assigned. Composite severity scores were calculated by summing grades across all lumbar levels for each pathology subtype, yielding pathology specific severity scores ranging from 0 to 30 for spinal stenosis, 0 to 40 for disc related pathologies, 0 to 20 for spondylolisthesis and 0 to 10 for facet arthropathy. An overall anatomical burden score was computed by summing category-level scores across all pathology domains, resulting in a total score ranging from 0 to 100.

### 2.4 Statistical analysis

#### 2.4.1 Biopsychosocial cLBP model

To evaluate the association between demographic and anthropometric, socioeconomic, psychological, anatomical determinants and the presence of cLBP, we developed a multivariable logistic regression model with cLBP presence as the binary outcome. Predictors included all four determinant domains: demographic and anthropometric factors (age, sex, BMI and race/ethnicity), socioeconomic factors (census tract level income and education, insurance status), psychological factors (depression related and anxiety related diagnoses) and anatomical determinants derived from radiology reports (stenosis, disc, spondylolisthesis, facet), as reported in **Table 1**. Continuous predictors (age, BMI, and anatomical severity scores) were modeled as continuous variables and were compared using Welch’s t-test. Median household income and college graduation rate were standardized (mean=0, standard deviation=1) prior to modeling to facilitate numerical convergence and enable comparison of effect sizes across variables with different scales. Categorical predictors (sex, race/ethnicity, insurance status, psychological diagnoses) were modeled using indicator variables with the reference category set as the class with highest cardinality and were compared using chi-square tests. Starting from the full multivariable logistic regression model containing all four domains, we performed a leave-one-domain-out analysis refitting the model four times, each time removing one domain while retaining the other three. Each reduced model was compared with the full model using the change in the Akaike information criterion (ΔAIC) (46) and the Bayesian information criterion (ΔBIC) (47), together with a likelihood-ratio test whose degrees of freedom equaled the number of parameters removed. A larger ΔAIC or ΔBIC indicates a greater unique contribution of the omitted domain to model fit relative to the remaining domains (48).

**Table 1.** Summary of all the variables included in the study. Data are presented as mean ± standard deviation for continuous variables and n (%) for categorical variables. Neighborhood characteristics are census tract-level estimates. Imaging severity scores were derived from radiology reports and graded at each spinal level as 0 (absent), 1 (mild-moderate), or 2 (severe) and then summed across L1-S1. P-values compare chronic vs non-chronic LBP using Welch’s t-test (continuous) or χ^2^ tests (categorical). P<0.05 was considered significant.

| Variable | Overall<br>(N=1480) | cLBP diagnosis<br>(N=973) | No cLBP diagnosis<br>(N=507) | p |
| --- | --- | --- | --- | --- |
| **Demographics and anthropometrics** |  |  |  |  |
| **Age** | 55.5 ± 16.2 | 54.6 ± 15.7 | 57.2 ± 17.0 | **0.005** |
| **Sex** |  |  |  | 0.242 |
| Male | 498 (34%) | 338 (35%) | 160 (32%) |  |
| Female | 982 (66%) | 635 (65%) | 347 (68%) |  |
| **BMI (kg/m<sup>2</sup>)** | 31.8 ± 8.1 | 32.0 ± 8.2 | 31.4 ± 7.97 | 0.158 |
| **Race/Ethnicity** |  |  |  | 0.220 |
| Non-Hispanic Black | 615 (42%) | 401 (41%) | 224 (42%) |  |
| Non-Hispanic White | 445 (30%) | 284 (29%) | 161 (32%) |  |
| Hispanic | 318 (21%) | 212 (22%) | 106 (21%) |  |
| Other | 102 (7%) | 76 (8%) | 26 (5%) |  |
| **Socioeconomic factors** |  |  |  |  |
| **Insurance status** |  |  |  | **0.036** |
| Private | 813 (55%) | 554 (57%) | 259 (51%) |  |
| Public | 667 (45%) | 419 (43%) | 248 (49%) |  |
| **Neighborhood characteristics** |  |  |  |  |
| Median household income (USD) | 77,413 ± 39,836 | 77,021 ± 40,468 | 78,166 ± 38,622 | 0.595 |
| College graduation rate (%) | 40.3 ± 27.3 | 40.3 ± 27.4 | 40.4 ± 27.2 | 0.961 |
| **Psychological factors** |  |  |  |  |
| Depression related diagnosis | 394 (27%) | 329 (34%) | 65 (13%) | **<0.001** |
| Anxiety related diagnosis | 552 (37%) | 442 (45%) | 110 (22%) | **<0.001** |
| **Anatomical factors** |  |  |  |  |
| Stenosis score | 4.32 ± 3.40 | 4.53 ± 3.41 | 3.91 ± 3.34 | **<0.001** |
| Disc score | 3.89 ± 2.34 | 4.02 ± 2.31 | 3.64 ± 2.37 | **0.004** |
| Listhesis score | 0.85 ± 1.07 | 0.91 ± 1.09 | 0.73 ± 1.02 | **0.002** |
| Facet arthropathy score | 3.08 ± 1.94 | 3.19 ± 1.85 | 2.87 ± 2.08 | **0.003** |
| Overall pathology score | 12.14 ± 7.14 | 12.65 ± 7.03 | 11.16 ± 7.26 | **<0.001** |

#### 2.4.2 Anatomical determinants model

To characterize associations between patient characteristics and imaging-derived pathology burden, we conducted secondary analyses modeling overall pathology score. Multivariable linear regression was performed with this composite score as the outcome variable and demographic and anthropometric, socioeconomic, and psychological factors as predictors as described above. Statistical significance was set at p<0.05. All analyses were performed using Python 3.11.5 with the statsmodels library (version 0.14.5).

## 3. Results

A total of 4,671 patients underwent lumbar spine MRI at Rush University System for Health between March 2014 and March 2024. After applying inclusion criteria, the final cohort comprised 1,480 patients (**Figure 1**). The cohort had a mean age of 55.5±16.2 years and included 982 (66%) female patients (**Table 1**). The population as predominantly Non-Hispanic Black (42%) or Non-Hispanic White (30%). Mean BMI was 31.8±8.1 kg/m^2^. Insurance coverage was 55% private and 45% public. Community-level socioeconomic characteristics included median household income of $77,413±$39,836 and college graduation rate of 40.3%±27.3%. Overall, 552 (37%) patients had a diagnosis of anxiety and 394 (27%) a diagnosis of depression. cLBP was diagnosed in 973 (66%) patients. Imaging-derived anatomical scores demonstrated the following pathology burden: 4.32±3.40 for stenosis, 3.89±2.34 for disc pathology, 3.08±1.94 for facet arthropathy, 0.85±1.07 for spondylolisthesis and 12.14±7.14 for the overall score comprising all pathologies. Compared with patients without cLBP, those with cLBP were younger and had higher stenosis, disc, facet, listhesis, and overall imaging severity scores, as well as higher rates of depression, anxiety, and private insurance (p-value<0.05) as shown in **Table 1**. BMI, census tract income and education, sex, and race/ethnicity did not differ between groups.

The LLM-based pipeline outperformed a simpler regex approach, achieving an overall F1-score of 0.98 compared to 0.92 for regex. Pathology-level results for the LLM pipeline and regex are summarized in **Table 2**. In multivariable logistic regression (**Table 3**), several factors were associated with cLBP diagnosis. Among demographics, older age was associated with lower odds of LBP diagnosis (OR=0.98, p<0.001), while male sex was associated with higher odds of diagnosis (OR=1.37, p=0.016). Regarding psychological factors, both depression (OR=2.69, p<0.001) and anxiety (OR= 2.31, p<0.001) were strongly associated with cLBP diagnosis. Among anatomical findings, higher stenosis (OR=1.07, p=0.006) and spondylolisthesis scores (OR=1.15, p=0.039) were associated with cLBP diagnosis. With respect to socioeconomic factors, public insurance coverage was associated with lower odds of LBP diagnosis (OR=0.69, p=0.003). No significant associations were observed for race/ethnicity and census tract-level socioeconomic factors.

**Table 2.** Comparison between regex based and NLP based pipeline.

| Pathology | LLM |  |  | Regex |  |  |
| --- | --- | --- | --- | --- | --- | --- |
|  | Recall | Precision | F1-score | Recall | Precision | F1-score |
| Foraminal stenosis | 0.89 | 1.0 | 0.92 | 0.75 | 1.0 | 0.86 |
| Central canal stenosis | 0.94 | 1.0 | 0.98 | 0.75 | 1.0 | 0.86 |
| Lateral recess stenosis | 0.96 | 1.0 | 0.98 | 0.85 | 1.0 | 0.92 |
| Disc bulging and herniation | 0.91 | 1.0 | 0.95 | 0.90 | 1.0 | 0.95 |
| Disc narrowing | 0.97 | 1.0 | 0.97 | 0.89 | 1.0 | 0.94 |
| Disc desiccation | 1.00 | 1.0 | 1.00 | 0.86 | 1.0 | 0.93 |
| Disc degeneration | 1.00 | 1.0 | 1.00 | 0.88 | 1.0 | 0.93 |
| Anterolisthesis | 1.00 | 1.0 | 1.00 | 0.98 | 1.0 | 0.99 |
| Retrolisthesis | 0.98 | 1.0 | 0.99 | 0.84 | 1.0 | 0.91 |
| Facet arthropathy | 0.94 | 1.0 | 0.97 | 0.79 | 1.0 | 0.88 |

**Table 3.** Results from multivariable logistic regression for LBP diagnosis, including odds ratios (OR) with 95% confidence intervals (CI) and p-values. Reference categories were set as the class with highest cardinality for each attribute. Income and education variables were standardized (mean=0, SD=1) prior to modeling to facilitate model convergence.

| Variable | OR | 95% CI | p-value |
| --- | --- | --- | --- |
| **Demographics and anthropometrics** |  |  |  |
| Age (per year) | 0.98 | 0.97 - 0.99 | **<0.001** |
| Sex (ref: Female) |  |  |  |
| Male | 1.37 | 1.06 - 1.76 | **0.016** |
| BMI (per kg/m<sup>2</sup>) | 1.00 | 0.99 - 1.02 | 0.732 |
| Race/Ethnicity (ref: Non-Hispanic Black) |  |  |  |
| Non-Hispanic White | 0.83 | 0.58 - 1.18 | 0.291 |
| Hispanic | 0.98 | 0.70 - 1.35 | 0.878 |
| Other | 1.65 | 0.98 - 2.79 | 0.060 |
| **Socioeconomic factors** |  |  |  |
| Insurance status (ref: Private) |  |  |  |
| Public | 0.69 | 0.54 - 0.88 | **0.003** |
| Neighborhood characteristics (standardized) |  |  |  |
| Median household income | 0.98 | 0.81 - 1.18 | 0.814 |
| College graduation rate | 0.99 | 0.81 - 1.20 | 0.905 |
| **Psychological factors** |  |  |  |
| Depression related diagnosis | 2.69 | 1.93 - 3.74 | **<0.001** |
| Anxiety related diagnosis | 2.31 | 1.74 - 3.05 | **<0.001** |
| **Anatomical factors (per point)** |  |  |  |
| Stenosis score | 1.07 | 1.02 - 1.13 | **0.006** |
| Disc score | 1.03 | 0.97 - 1.10 | 0.335 |
| Listhesis score | 1.15 | 1.00 - 1.31 | **0.039** |
| Facet arthropathy score | 1.08 | 0.99 - 1.18 | 0.082 |

The relative contribution of each domain (demographics and anthropometrics; socioeconomic; psychological; anatomical) to LBP diagnosis is summarized in **Table 4**. All four domains contributed significantly (likelihood-ratio test, all p<0.05), but their contributions differed substantially. Psychological factors accounted for the largest share of model fit (ΔAIC=120.1, ΔBIC=109.5, χ^2^(2)=124.1, p<0.001), followed by anatomical factors (ΔAIC=37.0, ΔBIC=15.8, χ^2^(4)=45.0, p<0.001). Demographics and anthropometrics showed a more limited contribution (ΔAIC=21.8, ΔBIC=-10.0, χ^2^(6)=33.8, p<0.001), as well as socioeconomic factors (ΔAIC=3.0, ΔBIC=-12.9, χ^2^(3)=9.0, p=0.029).

**Table 4.**
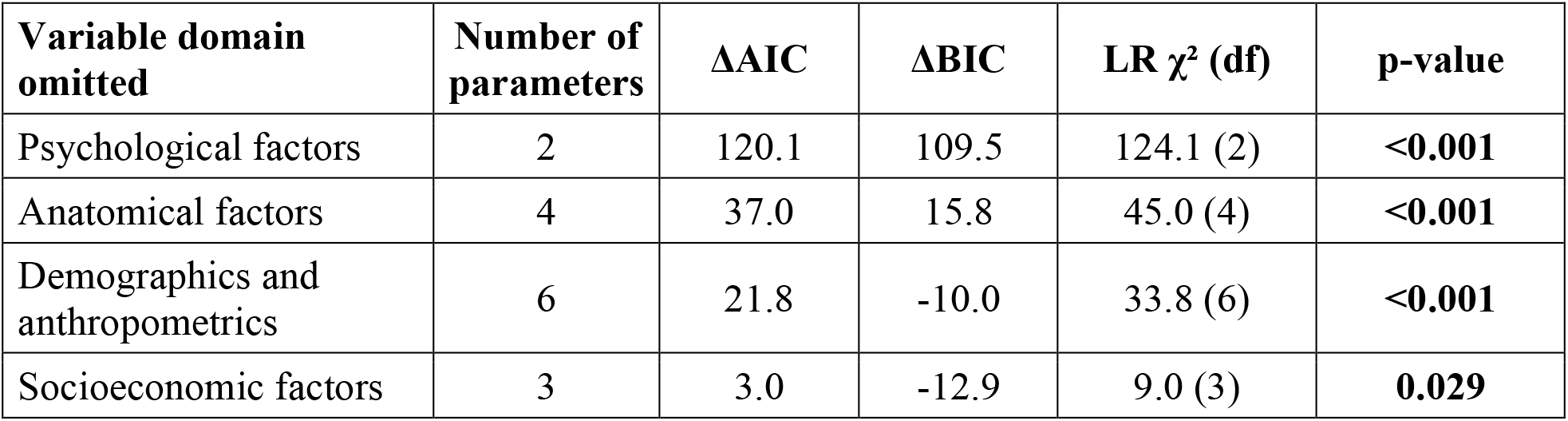
Relative contribution of each variable domain to the chronic low back pain model, assessed by leave-one-domain-out comparison against the full multivariable logistic regression model.

Greater anatomical burden was significantly associated with demographic characteristics and anthropometrics, including older age (β=0.26, p<0.001), male sex (β=1.58, p<0.001), higher BMI (β = 0.09, p<0.001), and Non-Hispanic White race compared with Non-Hispanic Black individuals (β = 0.98, p = 0.032) (**Table 5**). In contrast, none of the socioeconomic or psychological factors were significantly associated with overall anatomical severity. Results from pathology-specific models are reported in the Supplementary Materials (**Tables S1-S4**).

**Table 5.** Results from multivariable linear regression predicting overall pathology score, calculated as the sum of individual pathology scores (stenosis, disc pathology, facet arthropathy and spondylolisthesis; range 0-100). Regression coefficients (β) with 95% confidence intervals (CI) and p-values are shown. Reference categories were set as the class with highest cardinality for each attribute. Income and education variables were standardized (mean=0, SD=1) prior to modeling to facilitate model convergence.

| Variable | $\beta$ | 95% CI | p-value |
| --- | --- | --- | --- |
| **Demographics and anthropometrics** |  |  |  |
| Age (per year) | 0.26 | 0.24 - 0.28 | <0.001 |
| Sex (ref: Female) |  |  |  |
| Male | 1.58 | 0.94 - 2.22 | <0.001 |
| BMI ( per kg/m<sup>2</sup>) | 0.09 | 0.05 - 0.13 | <0.001 |
| Race/Ethnicity (ref: Non-Hispanic Black) |  |  |  |
| Non-Hispanic White | 0.98 | 0.08 - 1.88 | 0.032 |
| Hispanic | 0.17 | -0.67 - 1.00 | 0.697 |
| Other | 0.77 | -0.51 - 2.05 | 0.240 |
| **Socioeconomic factors** |  |  |  |
| Insurance status (ref: Private) |  |  |  |
| Public | -0.23 | -0.86 - 0.41 | 0.480 |
| Neighborhood characteristics (standardized) |  |  |  |
| Median household income | -0.02 | -0.52 - 0.48 | 0.935 |
| College graduation rate | 0.21 | -0.29 - 0.71 | 0.411 |
| **Psychological factors** |  |  |  |
| Depression related diagnosis | 0.43 | -0.34 - 1.29 | 0.272 |
| Anxiety related diagnosis | 0.10 | -0.60 - 0.79 | 0.790 |

## 4. Discussion

This study jointly examined demographic, anthropometric, socioeconomic, psychological and anatomical determinants of cLBP in a Chicago-based cohort of 1,480 patients undergoing lumbar spine MRI. Greater severity of spinal stenosis and listhesis were each associated with cLBP, with psychological comorbidities, including depression and anxiety, also showing strong associations. In contrast, pathology burden was associated with demographic and anthropometric factors, including age, sex, BMI, and race/ethnicity, and was not significantly associated with socioeconomic or psychological variables. Together, these findings demonstrate a dissociation between the factors associated with structural spinal degeneration and those associated with cLBP diagnosis, highlighting the multifactorial and biopsychosocial nature of LBP.

Consistent with prior work, imaging findings of spinal stenosis and listhesis were associated with higher odds of LBP diagnosis, supporting a relationship between these degenerative features and pain (49) (50). However, these associations were modest, reinforcing that structural abnormalities alone do not explain pain (12) (51). In contrast, anxiety and depression were strongly correlated with increased odds of LBP, aligning with extensive literature describing bidirectional relationships between chronic pain and psychological factors (41) (42). Additionally, we found that when removing psychological factors from the model, the model’s ability to distinguish patients with and without cLBP dropped the most, suggesting that psychological factors have unique information about cLBP that other domains do not contain. Psychological comorbidity may influence pain perception and healthcare seeking behavior, and its association with cLBP in this cohort aligns with the psychosocial drivers of pain beyond imaging findings.

Associations between cLBP and demographic, anthropometric, and socioeconomic status were also explored. Older age was associated with lower odds of cLBP diagnosis (**Table 3**) despite greater anatomical degeneration with advancing age (**Table 5**) (52) (53). While socioeconomic factors have been reported to be associated with LBP in population studies (54) (30) (55), the absence of such correlation in this cohort suggests that diagnostic patterns may be influenced more by clinical and psychological factors than by socioeconomic context. Male sex was associated with higher odds of LBP diagnosis, while public insurance coverage was associated with lower odds of a recorded diagnosis. These findings contrast with the broader literature, where LBP prevalence is higher among women and individuals with lower socioeconomic status (5) (56). As such, both associations should be interpreted with caution. Associations with sex may reflect differences in referral and management patterns in spine care and influence which patients enter an MRI-based cohort (57), rather than a purely biologic difference. Likewise, lower risk among publicly insured patients may reflect differences in access to specialty spine care and follow-up, which could reduce the likelihood that LBP is documented within the care pathway (58) (59). Taken together, these findings may capture healthcare access in an MRI-based EHR cohort rather than true differences in underlying disease prevalence.

The domain level leave-one-out comparison reinforced the primary analysis as psychological factors were stronger contributor to cLBP relative to the other domains, including anatomical factors. Demographic and socioeconomic factors improved model fit under the AIC but not the BIC, which favored the model without them. Because the BIC penalizes complexity more heavily, this indicates that both the demographic and socioeconomic domains carried predictive information without a parsimonious contribution.

In contrast to the LBP diagnosis model, overall anatomical pathology burden demonstrated strong associations with demographic characteristics. Older age, male sex, higher BMI, and Non-Hispanic White race were associated with greater degenerative changes, consistent with established biological and biomechanical processes underlying spinal degeneration (51) (60) (61). Neither socioeconomic or psychological variables were associated with overall anatomical severity. This finding suggests that while psychological factors may influence pain experience, they do not appear to drive the degenerative changes observed on imaging. This highlight a disconnect between factors associated with imaging findings and those associated with LBP diagnosis, reinforcing that structural pathology and pain are related but distinct constructs.

When examining specific severity scores, i.e. stenosis, disc, facet arthropathy, and listhesis, age was consistently associated with greater pathology across all four, in line with the well-established role of aging in spinal degeneration (51). Sex, BMI, and race/ethnicity showed more selective patterns: male sex was associated with higher stenosis and disc scores, higher BMI with greater stenosis, disc and facet arthropathy scores, while both Non-Hispanic White and Hispanic patients exhibited higher listhesis scores. These findings mirror the associations observed for overall anatomical severity.

The comparison between extraction strategies from radiology reports highlights a precision-recall trade-off between regex-based and LLM-based approaches. Regex achieved perfect precision across all pathologies, indicating that extracted cases are highly reliable and free of false positives. Thus, regex is well-suited for large-scale screening tasks where the objective is to identify a highly specific cohort, even at the cost of increased false negatives. Regex recall was indeed lower than LLM recall, reflecting its inability to capture the variability of radiology language. In contrast, the LLM-based pipeline maintains high precision and recall, enabling a robust characterization of disease burden, likely due to context-aware parsing as opposed to the deterministic pattern matching of regex. These benefits must be balanced against practical considerations as LLMs introduce higher computational cost, often requiring GPU resources and could exhibit non-deterministic behavior across runs. Consequently, regex remains advantageous for targeted, high-precision extraction, whereas LLMs are better suited for analyses where completeness and recall are critical. This pattern of high-precision/lower-recall regex versus more balanced LLM performance is consistent with prior work. A recent systematic review of natural language processing (NLP) methods for clinical registry population reported that rule-based approaches consistently achieved high precision but substantially lower recall compared to machine learning based methods, largely due to their inability to capture linguistic variability without exhaustive rule curation (62).

Some limitations should be acknowledged. First, the study was conducted at a single academic medical center in a population selected for MRI. This introduces geographic and referral bias, such as imaging performed for reasons unrelated to LBP, likely influencing LBP and pathology prevalence. Prospective population-based studies are necessary to confirm our findings. Second, LBP was identified using diagnosis codes, which may not accurately reflect true symptom prevalence, as diagnosis depends on both patient-reported symptoms, patient-physician interactions, and physician documentation practices. Third, patients without health insurance were not included in the analysis due to the very small number of uninsured patients in the cohort, limiting the ability to assess associations within this group. Finally, factors such as occupational exposures, physical activity, and treatment history were not accounted for and may contribute to confounding.

## 5. Conclusions

This study evaluates the relative contributions of demographic, anthropometric, socioeconomic, psychological and anatomical determinants of cLBP within a unified framework. In addition, to contextualize anatomical findings within the biopsychosocial model, associations between demographic, anthropometric, socioeconomic, psychological and anatomical factors were assessed. Our findings indicated that cLBP was primarily associated with psychological comorbidities and spinal pathologies, while spinal pathologies showed associations with age, sex, BMI and race/ethnicity. Next steps include a longitudinal approach to better characterize temporal relationships between imaging findings and pain. Additionally, extending this work to multi-institutional datasets could strengthen the validity of the findings.

## Data Availability

All data produced in the present study are available upon reasonable request to the authors.

## Statements and Declarations

This work was supported by the National Institutes of Health (NIH) under Grant R00AR077685. N.J.L. reports consulting fees from Medtronic; T.J.J. reports consulting fees from Medela AG and Tanoma Consulting, honoraria from Medela AG, royalties from the American College of Healthcare Executives, payment for expert testimony, grant-review honoraria from the Donaghue Foundation and AHRQ, research grants to her institution, and an unpaid leadership role with the International Society for Research in Human Milk and Lactation. The remaining authors declare no conflicts of interest.

## Bibliography

1. Hoy D, Brooks P, Blyth F, Buchbinder R. The Epidemiology of low back pain. Best Pract Res Clin Rheumatol [Internet]. 2010;24(6):769–81. Available from: 10.1016/j.berh.2010.10.002

2. Dagenais S, Caro J, Haldeman S. A systematic review of low back pain cost of illness studies in the United States and internationally. Spine J. 2008;8(1):8–20.

3. Itz CJ, Geurts JW, Van Kleef M, Nelemans P. Clinical course of non-specific low back pain:A systematic review of prospective cohort studies set in primary care. Eur J Pain. 2013;17(1):5–15.

4. Hestbaek L, Leboeuf-Yde C, Manniche C. Low back pain:what is the long-term course ? A review of studies of general patient populations. Eur Spine J. 2003;12(2):149–65.

5. Hartvigsen J, Hancock MJ, Kongsted A, Louw Q, Ferreira ML, Genevay S, et al. What low back pain is and why we need to pay attention. Lancet. 2018;391(10137):2356–2367.

6. Foster NE, Anema JR, Cherkin D, Chou R, Cohen SP, Gross DP, et al. Prevention and treatment of low back pain: evidence, challenges, and promising directions. Lancet. 2018;391(10137):2368–2383.

7. Pons E, Braun L, Hunink M, Kors J. Natural Language Processing in Radiology: A Systematic Review. Radiology. 2016;279(2):329–43.

8. Casey A, Davidson E, Poon M, Dong H, Duma D, Grivas A, et al. A systematic review of natural language processing applied to radiology reports. BMC Med Inform Decis Mak [Internet]. 2021;21(1):179. Available from: 10.1186/s12911-021-01533-7

9. Tan WK, Hassanpour S, Heagerty PJ, Rundell SD, Suri P, Huhdanpaa HT, et al. Comparison of Natural Language Processing Rules-based and Machine-learning Systems to Identify Lumbar Spine Imaging Findings Related to Low Back Pain. Acad Radiol [Internet]. 2018;25(11):1422–32. Available from: 10.1016/j.acra.2018.03.008

10. Wang Y, Sohn S, Liu S, Shen F, Wang L, Atkinson EJ, et al. A clinical text classification paradigm using weak supervision and deep representation. BMC Med Inform Decis Mak. 2019;19(1):1.

11. Ziegeler K, Kreutzinger V, Tong MW, Chin CT, Bahroos E, Wu PH, et al. Information Extraction from Lumbar Spine MRI Radiology Reports Using GPT4:Accuracy and Benchmarking Against Research-Grade Comprehensive Scoring. Diagnostics. 2025;15(7):930.

12. Brinjikji W, Diehn FE, Jarvik JG, Carr CM, Kallmes DF, Murad MH, et al. MRI findings of disc degeneration are more prevalent in adults with low back pain than in asymptomatic controls: A systematic review and meta-analysis. Am J Neuroradiol. 2015;36(12):2394–9.

13. Herlin, C; Kjaer, P; Espeland, A; Skouen, JS; Leboeuf-Yde, C; Karppinen, J; Niinimäki, J; Sørensen, JS; Storheim, K; Jensen T. Modic changes — Their associations with low back pain and activity limitation:A systematic literature review and meta-analysis. PLoS One. 2018;13(8):e0200677.

14. Yang D, Becker L, Hoehl BU, Mödl L, Zhang T, Liu S, et al. Association between MRI findings of lumbar morphometric changes and the characteristics of low back pain, pain-related disability, and quality of life: a cross-sectional study. Acad Radiol [Internet]. 2025;32(10):6000–8. Available from: 10.1016/j.acra.2025.06.026

15. Singh, Roop; Kumar, Pradeep; Wadhwani, Jitendra; Yadav, Rohtas K; Khanna, Mohit; Kaur S. A comparative study to evaluate disc degeneration on magnetic resonance imaging in patients with chronic low back pain and asymptomatic individuals. J Orthop Trauma Rehabil. 2021;28:1–7.

16. Kasch R, Truthmann J, Hancock MJ, Maher CG, Otto M, Nell C, et al. Association of Lumbar MRI Findings with Current and Future Back Pain in a Population-based Cohort Study. Spine (Phila Pa 1976). 2022;47(3):201–11.

17. Kalichman L, Cole R, Kim DH, Li L, Suri P, Guermazi A, et al. Spinal stenosis prevalence and association with symptoms:the Framingham Study. Spine J [Internet]. 2009;9(7):545–50. Available from: 10.1016/j.spinee.2009.03.005

18. Ziegeler K, Gensler LS, Link TM, Roach C, Scheffler AW, Bonnheim N, et al. Association between MRI findings and inflammatory symptoms in non-specific chronic low back pain. Eur Spine J. 2025;34(12):5530–8.

19. Han CS, Maher CG, Steffens D, Diwan A, Magnussen J, Hancock EC, et al. Some magnetic resonance imaging findings may predict future low back pain and disability:a systematic review. J Physiother [Internet]. 2023;69(2):79–92. Available from: 10.1016/j.jphys.2023.02.007

20. Jensen M, Brant-Zawadzki M, Obuchowski N, Modic M, Malkasian D, Ross J. Magnetic Resonance Imaging of the Lumbar Spine in People without Back Pain. N Engl J Med. 1994;331(2):69–73.

21. Depalma MJ, Ketchum JM, Saullo TR. Multivariable Analyses of the Relationships Between Age, Gender, and Body Mass Index and the Source of Chronic Low Back Pain. Pain Med. 2012;13(4):498–506.

22. Shmagel A, Foley R, Ibrahim H. Epidemiology of Chronic Low Back Pain in US Adults:Data From the 2009 – 2010 National Health and Nutrition Examination Survey. Arthritis Care Res Hoboken. 2016;68(11):1688–94.

23. Otero-ketterer E, Peñacoba-puente C, Ferreira Pinheiro-araujo C, Valera-calero JA, Ortega-santiago R. Biopsychosocial Factors for Chronicity in Individuals with Non-Specific Low Back Pain:An Umbrella Review. Int J Env Res Public Heal. 2022;19(16):10145.

24. Burke C, Fillipo R, Epplein M, Brookhart M, Bosworth HB, Goode AP. Racial and Ethnic Disparities in the Incidence and Prevalence of Low Back Pain in the United States: A Systematic Review. Arthritis Care Res Hoboken. 2025;78(1):141–53.

25. Carey TS, Garrett JM. The Relation of Race to Outcomes and the Use of Health Care Services for Acute Low Back Pain. Spine (Phila Pa 1976). 2003;28(4):390–4.

26. Green CR, Anderson KO, Baker TA, Campbell LC, Decker S, Fillingim RB, et al. The Unequal Burden of Pain:Confronting Racial and Ethnic Disparities in Pain. Pain Med. 2003;4(3):277–294.

27. Campbell CM, Edwards RR. Ethnic differences in pain and pain management. Pain Manag. 2012;2(3):219–30.

28. Roseen EJ, Smith CN, Essien UR, Cozier YC, Joyce C, Morone NE, et al. Racial and Ethnic Disparities in the Incidence of High-Impact Chronic Pain Among Primary Care Patients with Acute Low Back Pain:A Cohort Study. Pain Med. 2023;24(6):633–43.

29. Shraim M, Cifuentes M, Willetts JL, Marucci-Wellman H, Pransky G. Regional socioeconomic disparities in outcomes for workers with low back pain in the United States. Am J Ind Med. 2017;60(5):472–83.

30. Karran EL, Grant AR, Moseley GL. Low back pain and the social determinants of health:a systematic review and narrative synthesis. Pain. 2020;161(11):2476–93.

31. Gonzalez G, da Silva T, Avanzi M, Macedo G, Alves S, Indini L, et al. Low back pain prevalence in Sao Paulo, Brazil: A cross-sectional study. Braz J Phys Ther. 2021;25(6):837– 845.

32. Louw QA, Morris LD, Grimmer-somers K. The Prevalence of low back pain in Africa:a systematic review. BMC Musculoskelet Disord. 2007;8:105.

33. Barrero LH, Hsu Y, Terwedow H, Perry MJ, Dennerlein JT, Brain JD, et al. Prevalence and Physical Determinants of Low Back Pain in a Rural Chinese Population. Spine (Phila Pa 1976). 2006;31(23):2728–34.

34. Bernier Carney KM, Guite JW, Young EE, Starkweather AR. Investigating key predictors of persistent low back pain:A focus on psychological stress. Appl Nurs Res [Internet]. 2021;58:151406. Available from: 10.1016/j.apnr.2021.151406

35. Huang Z, Guo W, Martin JT. Socioeconomic status, mental health, and nutrition are the principal traits for low back pain phenotyping:Data from the osteoarthritis initiative. JOR Spine. 2023;6(2):e1248.

36. Kao Y, Chen J, Chen H, Liao K, Huang S. The association between depression and chronic lower back pain from disc degeneration and herniation of the lumbar spine. Int J Psychiatry Med. 2022;57(2):165–77.

37. Linton SJ. A Review of Psychological Risk Factors in Back and Neck Pain. Spine (Phila Pa 1976). 2000;25(9):1148–56.

38. Pincus T, Burton AK, Vogel S, Field AP. A Systematic Review of Psychological Factors as Predictors of Chronicity / Disability in Prospective Cohorts of Low Back Pain. Spine (Phila Pa 1976). 2002;27(5):e109–e120.

39. Almeida CS de, Miccoli LS, Andhini NF, Aranha S, Oliveira LC de, Artigo CE, et al. No 主観的健康感を中心とした在宅高齪者における健康関連指標に関する共分散構造分析 Title. Rev Bras Linguística Apl [Internet]. 2016;5(1):1689–99. Available from: https://revistas.ufrj.br/index.php/rce/article/download/1659/1508%0Ahttp://hipatiapress.com/hpjournals/index.php/qre/article/view/1348%5Cnhttp://www.tandfonline.com/doi/abs/10.1080/09500799708666915%5Cnhttps://mckinseyonsociety.com/downloads/reports/Educa

40. Jarvik JG, Hollingworth W, Heagerty PJ, Haynor DR, Boyko EJ, Deyo RA. Three-Year Incidence of Low Back Pain in an Initially Asymptomatic Cohort: Clinical and Imaging Risk Factors. Spine (Phila Pa 1976). 2005;30(13):1541–8.

41. Hooten WM. Chronic Pain and Mental Health Disorders: Shared Neural Mechanisms, Epidemiology, and Treatment. Mayo Clin Proc [Internet]. 2016;91(7):955–70. Available from: 10.1016/j.mayocp.2016.04.029

42. Kroenke K, Wu J, Bair MJ, Krebs EE, Damush TM, Tu W. Reciprocal Relationship Between Pain and Depression: A 12-Month Longitudinal Analysis in Primary Care. J Pain [Internet]. 2011;12(9):964–73. Available from: 10.1016/j.jpain.2011.03.003

43. Bishop-Royse J, Saiyed N, Schober D, Laflamme E, Lange-Maia B, Ferrera M, et al. Cause -Specific Mortality and Racial Differentials in Life Expectancy, Chicago 2018 – 2019. J Racial Ethn Heal Disparities. 2023;11(2):846–852.

44. Chicago Atlas. https://chicagohealthatlas.org/.

45. Adekkanattu P, Olfson M, Susser LC, Patra B, Vekaria V, Coombes BJ, et al. Comorbidity and healthcare utilization in patients with treatment resistant depression: A large-scale retrospective cohort analysis using electronic health records. J Affect Disord. 2023;324:102– 13.

46. Akaike H. A new look at the statistical model identification. IEEE Trans Automat Contr. 1974;19(6):716–23.

47. Schwarz G. Estimating the dimension of a model. Ann Stat. 1978;6(2):461–4.

48. Burnham KP, Anderson DR. Multimodel inference: understanding AIC and BIC in model selection. Sociol Methods Res. 2004;33(2):261–304.

49. Endean A, Palmer KT, Coggon D. Potential of MRI Findings to Refine Case Definition for Mechanical Low Back Pain in Epidemiological Studies: A Systematic Review. Spine (Phila Pa 1976). 2011;36(2):160–9.

50. De Schepper EIT, Damen J, Van Meurs JBJ, Ginai AZ, Popham M, Hofman A, et al. The association between lumbar disc degeneration and low back pain: The influence of age, gender, and individual radiographic features. Spine (Phila Pa 1976). 2010;35(5):531–6.

51. Brinjikji W, Luetmer P, Comstock B, Bresnahan B, Chen LE, Deyo RA, et al. Systematic Literature Review of Imaging Features of Spinal Degeneration in Asymptomatic Populations. AJNR Am J Neuroradiol. 2015;36(4):811–6.

52. Wong AYL, Karppinen J, Samartzis D. Low back pain in older adults:risk factors, management options and future directions. Scoliosis Spinal Disord. 2017;12:14.

53. Docking RE, Fleming J, Brayne C, Zhao J, Macfarlane GJ, Jones GT. Epidemiology of back pain in older adults:prevalence and risk factors for back pain onset. Rheumatology. 2011;50(9):1645–53.

54. Ikeda T, Sugiyama K, Aida J, Tsuboya T, Watabiki N, Kondo K, et al. Socioeconomic inequalities in low back pain among older people:the JAGES cross-sectional study. Int J Equity Heal. 2019;18(1):15.

55. Mathieu J, Roy K, Robert M-ève, Akeblersane M, Descarreaux M, Marchand A. Sociodemographic determinants of health inequities in low back pain:a narrative review. Front Public Heal. 2024;12:1392074.

56. Calais Ferreira L, Pozzobon D, Pinheiro MB, Blyth FM, Ordoñana JR, Duncan GE, et al. Sex differences in lifetime prevalence of low back pain:A multinational study of opposite--sex twin pairs. Eur J Pain. 2023;27(10):1150–60.

57. Taylor BA, Casas-ganem J, Vaccaro AR, Hilibrand AS, Hanscom B, Albert TJ. Differences in the Work-Up and Treatment of Conditions Associated With Low Back Pain by Patient Gender and Ethnic Background. Spine (Phila Pa 1976). 2005;30(3):359–64.

58. Felan NA, Burger EL, Lind DRG, Sidrak JP, Stokes DJ, Seawalt T, et al. Impact of Medicaid, Medicare, and private insurance on access to orthopedic surgeons of the spine:a national mystery caller study. Spine J [Internet]. 2026; In press. Available from: 10.1016/j.spinee.2025.12.006

59. Segal DN, Grabel ZJ, Shi WJ, Gottschalk MB, Boden SD. The impact of insurance coverage on access to orthopedic spine care. J Spine Surg. 2018;4(2):260–3.

60. Samartzis D, Karppinen J, Chan D, Luk KDK, Cheung KMC. The Association of Lumbar Intervertebral Disc Degeneration on Magnetic Resonance Imaging With Body Mass Index in Overweight and Obese Adults: A Population-Based Study. Arthritis Rheum. 2012;64(5):1488–96.

61. Lambrechts MJ, Pitchford C, Hogan D, Li J, Fogarty C, Rawat S, et al. Lumbar spine intervertebral disc desiccation is associated with medical comorbidities linked to systemic inflammation. Arch Orthop Trauma Surg [Internet]. 2023;143(3):1143–53. Available from: 10.1007/s00402-021-04194-3

62. Liu L, Blake V, Barman M, Gallego B, Churches T, Kennedy G, et al. Using natural language processing to extract information from clinical text in electronic medical records for populating clinical registries:a systematic review. JAMIA. 2026;33(2):484–99.s

